# Neutralizing response against E484K variant after original SARS-CoV-2 infection

**DOI:** 10.1101/2021.05.07.21256803

**Authors:** Takuma Hayashi, Nobuo Yaegashi, Ikuo Konishi

**Affiliations:** National Hospital Organization Kyoto Medical Center, Kyoto, Japan; START, Japan Science and Technology Agency (JST), Tokyo, Japan; Department of Obstetrics and Gynecology, Tohoku University School of Medicine, Miyagi, Japan; Department of Obstetrics and Gynecology, Kyoto University School of Medicine, Kyoto, Japan; Immediate Past President, Asian Society of Gynecologic Oncology, Tokyo, Japan

**Keywords:** **RBD** mutation, Neutralizing antibody, Spike glycoprotein, **SARS-CoV-2**, **ACE2**

## Abstract

Severe acute respiratory syndrome-coronavirus-2 (SARS-CoV-2) variants, which are spreading in the United Kingdom (UK) and elsewhere, have been found in infected individuals in Japan. The virus mutates, to facilitate its life in the host, during the process of repeated proliferation in the body of the host, including humans. In other words, it is natural that a human-compatible mutant strain always predominates in infection and proliferation. As a result, the viral mutants acquire strong proliferative potential in the host and are highly pathogenic. The number of people infected with the mutated SARS-CoV-2 variant E484K, which is different from the SARS-CoV-2 variants that are spreading in the UK, South Africa, and Brazil, is increasing in Tokyo. It has been pointed out that the effects of immunity and vaccines may be reduced against the Tokyo-type SARS-CoV-2 variant E484K. We have investigated the neutralization response to various mutations in the spike glycoprotein using the serum of people already infected with the original SARS-CoV-2. The results showed that SARS-CoV-2 variants with Y543F or N501Y mutations in the spike glycoprotein affect the neutralization reaction. However, single E484K mutations within the spiked glycoprotein of the Tokyo-type SARS-CoV-2 variant are unlikely to have a significant effect on the affinity of the host antibody for the virus.

## Introduction

According to a report from the World Health Organization (WHO), in the week of March 29–April 4, 2021, the number of new people infected with severe acute respiratory syndrome-coronavirus-2 (SARS-CoV-2) worldwide was more than 4 million. The number of new people infected with SARS-CoV-2 on a weekly basis had increased for the sixth straight week (1). The cause of the increase in the number of people newly infected with SARS-CoV-2 is that SARS-CoV-2 variants, which are said to be more efficient in infecting humans, are spreading all over the world. Infection with variants of SARS-CoV-2 is widespread and seen worldwide. People infected with the SARS-CoV-2 variant 20I/501Y.V1/B.1.1.7, called the British-type, have been confirmed in 109 countries/regions (2,3). Persons infected with the SARS-CoV-2 variant 20H/501Y.V2/B.1.351, called the South African type, have been confirmed in 73 countries/regions (4).

According to a report by the National Institute of Infectious Diseases in Japan, on March 3, 2021, at the quarantine center at Narita Airport (New Tokyo International Airport), the SARS-CoV-2 variant E484K was confirmed in two Japanese individuals. Until March 31, 2021, 394 cases infected with the Tokyo-type SARS-CoV-2 variant E484K were confirmed in Japan. Perhaps the SARS-CoV-2 variant E484K came to Japan from abroad, and hence its infection is spreading in the Tokyo metropolitan area. However, it is possible that the original SARS-CoV-2 has mutated to E484K in Japan. In the Tokyo metropolitan area, the SARS-CoV-2 variant E484K has been detected in one-third of all infected individuals confirmed to be positive for SARS-CoV-2 by reverse transcription-polymerase chain reaction (RT-PCR) (5). The coronavirus disease 2019 (COVID-19) research group in Tokyo reported that the Tokyo-type SARS-CoV-2 variant E484K has replaced the original SARS-CoV-2 as the causative virus of COVID-19 in the Tokyo metropolitan area.

It has been pointed out that SARS-CoV-2 variants may acquire the property of escaping the attack of neutralizing antibodies when mutations occur in the spike glycoprotein of SARS-CoV-2 (6,7,8). Therefore, it has been pointed out that patients with SARS-CoV-2 may be easily reinfected and that the antiviral effect of the COVID-19 vaccine may not be observed (6,7,8,9). We therefore investigated the affinity of immunoglobulin G (IgG) for the original receptor binding domain (RBD) and the mutant RBDs—Y453F, N501Y, and E484K—in the serum samples obtained from 41 COVID-19-positive patients and 20 COVID-19-negative patients. The findings indicate that SARS-CoV-2 infection with Y453F, N501Y, or E484K mutations in the spike glycoprotein of SARS-CoV-2 may escape the antiviral effect of neutralizing antibodies or COVID-19 vaccination.

## Materials and Methods

### Samples

As soon as a resident developed COVID-19, the testing recommendations from the Japan Ministry of Health, Labor, and Welfare (Chiyoda, Tokyo, Japan) and Tokyo Metropolitan Research Center for Health and Safety (Shinjuku, Tokyo, Japan) were followed, and all participants were repeatedly tested using Reverse Transcription-Polymerase Chain Reaction (RT-PCR) test on nasopharyngeal swabs or an anti-SARS-CoV-2 IgG/IgM test until no new cases were diagnosed. Participants provided written informed consent, and the study was approved by the Japan National Hospital Organization Central Ethics Review Board for Clinical Research (Meguro, Tokyo, Japan).

In participants with or without a prior history of COVID-19, we compared RT-PCR test or IgG/IgM antibody levels against SARS-CoV-2 proteins S and N by using two-sided Wilcoxon Mann-Whitney tests. The statistical significance threshold was set at 5%. Analyses were performed using XLSTAT version 2021.2 (Addinsoft &Vose Software, NY, USA).

### Quantitative measurement of SARS-CoV-2 neutralizing antibodies

Participants were randomly contacted based on stratification into two groups: RT-PCR-positive or anti-SARS-CoV-2 IgG/IgM-positive with history of symptomatic COVID-19 (Group I) and asymptomatic COVID-19 and RT-PCR or anti-SARS-CoV-2 IgG/IgM-negative people (Group II). Plasma was tested using enzyme-linked immunosorbent assay (ELISA) for IgG to RBD of spike trimer or RBD variants of spike trimer, which was modified from an assay (10) to give a readout of half-maximal binding titers by using The SeroFlash™ SARS-CoV-2 Neutralizing Antibody Assay Fast Kit (EpiGentek Group Inc., NY, USA) (details are shown in Supplementary Appendix). Samples were compared between Group I and Group II.

### Analysis of the three-dimensional structures of RBD of spike glycoprotein of SARS-CoV-2 with neutralizing antibodies or the binding sites of human angiotensin-converting enzyme 2 (ACE2)

Data on the three-dimensional structure of the RBD of the spike glycoprotein of original SARS-CoV-2 PDB ENTITY SEQ 6VW1_1 and SARS-CoV-2 variants were used in these studies (5). Data (PDB: 6XC2, 6XC4, 7JMP, 7JMO, 6XKQ, 6XKP, and 6XGD) on the three-dimensional structure of six neutralizing antibodies (CC12.1, CC12.3, COVA2-39, COVA2-04, CV07-250, CV07-270) and the REGN-COV2 (REGN10933 and REGN10987) antibodies that bind to the spike glycoprotein of original SARS-CoV-2 or SARS-COV-2 variants were used in these studies (11).

Using the Spanner program, we predicted the three-dimensional structure of the SARS-CoV-2 spike glycoprotein Y453F mutant based on PDB (ENTITY SEQ 6VW1_1). We investigated the binding of the spike glycoprotein Y453F mutant of SARS-CoV-2 to human ACE2 and determined the affinity of Y453F, N501Y, and E484K mutants of the spike glycoprotein of SARS-CoV-2 to six neutralizing monoclonal antibodies using the MOE program (three-dimensional protein structure modeling, protein-protein docking analysis: MOLSIS Inc., Tokyo, Japan) and Cn3D macromolecular structure viewer. We identified the epitope regions within the molecule of the spike glycoprotein of original SARS-CoV-2 and SARS-CoV-2 E484K variant by using the Immune Epitope Database (IEDB) (http://www.iedb.org/home_v3.php).

(Details regarding the three-dimensional structures modeling and a microneutralization assay are provided in the Supplementary Appendix, available with the full text of this article at XXX.)

## Results

### Structural analysis

The results from the Spanner analysis revealed that the RBD mutants Y453F, N501Y, and E484K did not affect the three-dimensional structure of the original SARS-CoV-2 spike glycoproteins (Supplementary Figure 1, Supplementary Figure 2). The results clarified that the affinity of the E484K mutant spike glycoprotein to human ACE2 was slightly higher than that of the original SARS-CoV-2 spike glycoprotein (Table 1, Supplementary Figure 1, Supplementary Figure 3A). Conversely, the affinity of the RBD E484K mutant of spike glycoprotein to human ACE2 was stronger than that of the original RBD of spike glycoprotein (Figure 1, Table 1). The slightly higher affinity observed in E484K was due to the replacement of Glu at position 484 by Lys, which was able to form a bond with the benzene ring with Phe at position 490 in RBD of spike glycoprotein (Supplementary Figure 3A). As a result, the RBD structure of the spiked glycoprotein becomes more stable. However, like the original RBD, the E484K mutant does not form a hydrogen bond with Lys at position 31 of the human ACE2 located near Lys 484 (Supplementary Figure 1, Supplementary Figure 3A). Taken together, the RBD F484K mutation is considered to have a non-significant effect on the affinity of spiked glycoproteins toward human ACE2.

**Table 1.** Affinity of conventional RBD and RBD of E484K mutant toward the neutralizing antibodies and serum from each patient. The binding of the original receptor binding domain (RBD) and RBD of E484K mutant to human angiotensin-converting enzyme 2 (ACE2) and to six neutralizing monoclonal antibodies was investigated using the MOE program and Cn3D macromolecular structure viewer. Binding energy, calculated by the MOE program, is shown in the table. Quantitative measurement of SARS-CoV-2 neutralizing antibody levels was examined using a SeroFlash™ SARS-CoV-2 Neutralizing Antibody Assay Fast Kit (EpiGentek Group Inc. NY, USA). A strong affinity as neutralizing antibody for the E484K mutant was shown for the serum IgG of 16 of the 41 COVID-19-positive patients. Moderate affinity as neutralizing antibody for the RBD E484K mutant was shown in the serum IgG of 10 of the 41 the COVID-19-positive patients. No affinity as a neutralizing antibody for the original RBD and the RBD E484K mutant was shown in the serum IgG of the COVID-19-negative subjects. Detailed experimental results are indicated in Supplementary Table 1.

| Binding energy between RBD and human ACE2 |  | Effect of mutation on antibody |
| --- | --- | --- |
| RBD | RBD (E484K) |  |
| -10.48 Kcal/mol | -10.89 Kcal/mol | Slightly higher affinity |

| Neutralizing antibody | Binding energy between RBD and antibodies |  | Effect of mutation on antibody |
| --- | --- | --- | --- |
|  | RBD | RBD (E484K) |  |
| CC12.1 | -12.49 Kcal/mol | -11.86 Kcal/mol | Slightly lower affinity |
| CC12.3 | -9.13 Kcal/mol | -9.95 Kcal/mol | Slightly higher affinity |
| COVA2-39 | -15.13 Kcal/mol | 13.21 Kcal/mol | Lower affinity |
| COVA2-04 | -9.73 Kcal/mol | -9.95 Kcal/mol | No effect |
| CV07-250 | -9.82 Kcal/mol | -10.05 Kcal/mol | No effect |
| CV07-270 | -14.91 Kcal/mol | -11.51 Kcal/mol | Lower affinity |
| REGN-COV2 | -15.32 Kcal/mol | 13.27 Kcal/mol | Lower affinity |

**Table 1. Affinity of conventional RBD and RBD of E484K mutant toward the neutralizing antibodies and serum from each patient.**
| Patient Serums | Number of samples with affinity for RBD |  | Effect of mutation on antibody |
| --- | --- | --- | --- |
|  | RBD | RBD (E484K) | E484K |
| Positive 41 samples | 29 samples | 26 samples | Slightly lower affinity |
| Negative 20 samples | 0 sample | 0 sample | NA |

| Binding energy between RBD and human ACE2 |  |  | Effect of mutation on antibody |
| --- | --- | --- | --- |
| RBD | RBD (E484K) |  |  |
| -10.48 Kcal/mol | -10.89 Kcal/mol |  | Slightly higher affinity |

**Figure 1.**
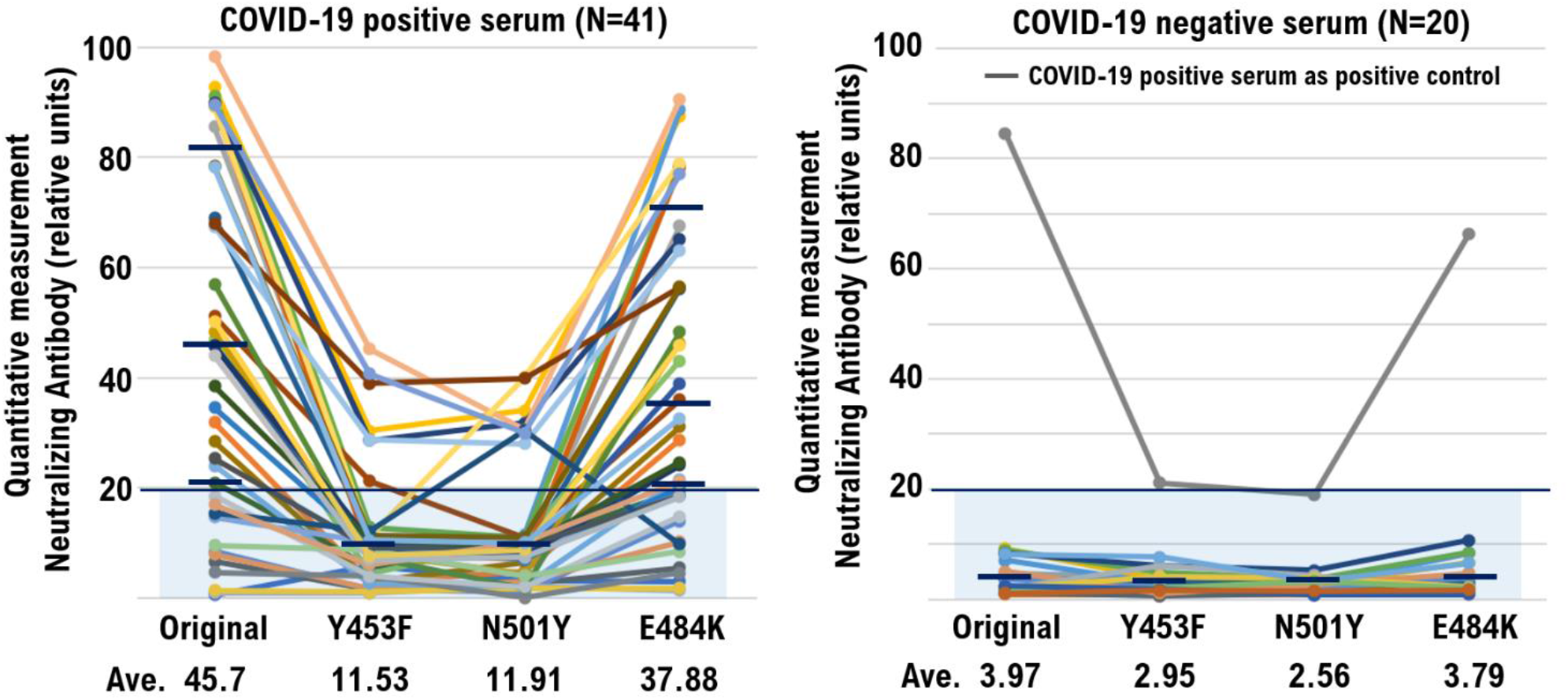
Neutralizing Response against the Original Virus and Variants after SARS-CoV-2 Infection. Serum samples from 41 patients previously infected with severe acute respiratory syndrome-coronavirus-2 (SARS-CoV-2), obtained 1 to 12 weeks after natural infection were tested with a microneutralization assay for the neutralizing response against sublineage of receptor binding domain (RBD) of the original virus and the RBD Y453F, RBD N501Y, and RBD E484K mutants. The lines at 20 indicate the cut-off titer. Solid lines and numbers indicate the geometric mean titer and error bars show 95% confidence intervals.

The results of the structural analysis revealed that the affinity between the RBD E484K mutant spike glycoprotein and three of the six examined monoclonal antibodies (CC12.1, CC12.3, COVA2-39, COVA2-04, CV07-250, CV07-270) was clearly weak compared with the original RBD (Table 1, Supplementary Figure 3B–3G, Supplementary Figure 3I). The results also demonstrated that the affinity between the RBD E484K mutant of spike glycoprotein and one of the six examined monoclonal antibody was clearly strong as compared to the original RBD (Table 1, Supplementary Figure 3C).

On September 28, 2020, the US pharmaceutical manufacturer Regeneron Pharmaceuticals announced the production of the antibody cocktail therapy, REGN-COV2, which combined two neutralizing antibodies, Casirivimab and Imdevimab, for the treatment and prevention of COVID-19 (9) (Figure 3A). On January 11, 2021, Regeneron Pharmaceuticals stated the likelihood of the antiviral effectiveness of REGN-COV2 on SARS-CoV-2 subspecies (12). Therefore, the affinity between REGN-COV2, used for antibody cocktail therapy, and the RBD E484K mutation in the spiked glycoprotein was investigated. The Glu at position 484 of original RBD of spike glycoprotein of SARS-CoV-2 forms hydrogen bonds with the Tyr at position 53, Ser at position 54, and Ser at position 56 of the REGN10933 antibody Fab fragment heavy chain, which is one of the components of REGN-COV2 (Table 1, Supplementary Figure 3H). Therefore, affinity between REGN-COV2 and original RBD of spike glycoprotein of SARS-CoV-2 is strong. However, in case of the RBD E484K mutant, the Lys at position 484 of RBD of spike glycoprotein of SARS-CoV-2 forms a hydrogen bond with only the Tyr at position 53 of the REGN10933 antibody Fab fragment heavy chain (Table 1, Supplementary Figure 3H). Based on these results, affinity between REGN-COV2 and the RBD E484K mutant of spike glycoprotein of SARS-CoV-2 is considered to become weak compared to original RBD of spike glycoprotein.

### Qualitative measurement of SARS-CoV-2 neutralizing antibodies

Next, we investigated the affinity of serum IgG as a neutralizing antibody toward the original RBD, and RBD E484K mutant in COVID-19-positive patients and COVID-19-negative subjects. A strong affinity as neutralizing antibody for the original RBD was shown by the serum IgG, in 29 of the 41 COVID-19-positive patients (Table 1, Supplementary Table 1). A strong affinity as neutralizing antibody for the E484K mutant was demonstrated by the serum IgG in 16 of the 41 COVID-19-positive patients (Table 1, Supplementary Table 1). Moderate affinity as neutralizing antibody for the RBD E484K mutant was shown by the serum IgG in 10 of the 41 COVID-19-positive patients (Figure 1A, Table 1, Supplementary Table 1). No affinity as neutralizing antibody for the original RBD, and the RBD E484K mutant was exhibited by the serum IgG of the COVID-19-negative subjects (Figure 1B, Table 1, Supplementary Table 1). Moderate affinity as neutralizing antibody for the RBD Y453F mutant was demonstrated by the serum IgG in 6 of the 41 COVID-19-positive patients (Table 1, Supplementary Table 1). Strong affinity as neutralizing antibody for the RBD Y453F mutant was shown by the serum IgG in 1 of the 41 COVID-19-positive patients (Table 1, Supplementary Table 1). Moderate affinity as neutralizing antibody for RBD N501Y mutant was shown by the serum IgG in 8 of the 41 COVID-19-positive patients (Table 1, Supplementary Table 1).

A recent report shows that the levels of antibodies were comparable between the deteriorated and stabilized groups in both males and females, but stabilized females tended to have higher antibody levels (13). In this experiment, the neutralizing antibody titer against the original RBD or RBD E484K mutant was higher in the IgG in the serum from infected males than infected females (Supplementary Table 1). However, this gender difference was not statistically significant. Frequencies of SARS-CoV-2 convergent clones were lower in adult blood and lymphoid tissues compared to children’s blood and lymphoid tissues (14). In this experiment, the antibody titer having neutralizing activity against the original RBD or RBD E484K mutant was lower in the sera from infected persons in their 20s and 30s than in other age groups (Supplementary Table 1). However, this age-specific difference was not statistically significant.

Based on these results, we concluded that the mutation of tyrosine (at amino acid residue 453) to phenylalanine and the mutation of asparagine (at amino acid residue 501) to tyrosine eliminated the inhibitory effects of the neutralizing antibody on binding between ACE2 and the RBD of the SARS-CoV-2 spike glycoprotein. It is possible that the affinity between the appropriate amino acid residues in the variable region of the antibody and the RBD of the Y453F and N501Y mutations was diminished owing to weak recognition of the monoclonal antibody by SARS-CoV-2 spike glycoproteins. Although a recent report speculated that mRNA vaccine BNT162b2-induced immune sera showed reduced inhibitory activity against viruses containing an E484K spike mutation (15), the single mutation of glutamic acid at amino acid residue 484 to ricin did not significantly prevent the inhibitory effects of the neutralizing antibody on binding between ACE2 and the RBD of the SARS-CoV-2 spike glycoprotein. Analysis using the IEDB does not elucidate the protein region including E484 as immune epitope (Supplementary Figure 4A and 4B). Taken together, the protein region including E484 might not play a role as the immune epitope.

## Discussion

So far, the SARS-CoV-2 variant 20H/501Y.V2/B.1.351 confirmed in South Africa and the SARS-CoV-2 variant P.1/20J/501Y.V3/B.1.1.248 confirmed in Brazil have been found to have N501Y and E484K mutations in the spike glycoprotein of SARS-CoV-2, which are thought to enhance the infectivity of the virus (16,17). Based on a report by the WHO, the infectivity of the SARS-CoV-2 variant 20I/501Y.V1/B.1.1.7 found in the UK is 36%–75% higher compared to the original SARS-CoV-2 (18). In addition, patients infected with SARS-CoV-2 variant B.1.1.7 are more likely to become severely ill. In people infected with this virus mutant, death risk is higher. The SARS-CoV-2 variant observed in South Africa (20H/501Y.V2/B.1.351) has both N501Y and E484K mutations. Compared to the original SARS-CoV-2, the infectivity of the SARS-CoV-2 variant B.1.351 is 50% higher. According to reports from South Africa, the mortality rate of people infected with the SARS-CoV-2 variant B.1.351 is 20% higher than in people infected with the original SARS-CoV-2. The SARS-CoV-2 variant P.1/20J/501Y.V3/B.1.1.248, which is widespread in Brazil, has both N501Y and E484K mutations. According to a report by the WHO, the infectivity of the SARS-CoV-2 variant B.1.1.248 is considered to be higher than that of the original SARS-CoV-2. However, individuals affected with SARS-CoV-2 variant B.1.1.248 do not necessarily suffer from severe COVID-19.

No N501Y mutations have been found in the spike glycoprotein of the new SARS-CoV-2 mutant, which is feared to have spread in the Tokyo metropolitan area, and only the E484K mutation has been found (19,20). The biological and medical properties of the Tokyo-type SARS-CoV-2 variant E484K have not been clarified. So, far, no aggravation of COVID-19 symptoms or enhancement of infectivity has been observed in people infected with Tokyo-type SARS-CoV-2 variant E484K. In addition, SARS-CoV-2 variants B.1.1.7, B.1.351, and B.1.1.248 are currently being detected in the screening of mutant variants conducted by local governments nationwide. In order to understand the biological and medical properties of the Tokyo-type SARS-CoV-2 variant E484K detected in serum obtained from infected people, the genetic information of the Tokyo-type SARS-CoV-2 variant E484K must be analyzed in detail.

The SARS-CoV-2 variant, which is spreading in the UK and other European countries, is being detected in many infected people in Japan. Normally, during the process of repeated proliferation in the living body of the host, including humans, the virus mutates to make it easier to live in the host. In other words, it is natural that a human-compatible mutant strain always predominates in infection and proliferation. As a result, the viral mutants acquire strong proliferative potential in the host and are highly pathogenic.

Inactivated vaccines have been the mainstream vaccines to date. However, the effectiveness of inactivated SARS-CoV-2 vaccines is unclear. The mRNA vaccine BNT162b2 is 90%–95% effective against SARS-CoV-2 infection. Vaccination cannot completely prevent viral infections, but it relieves the symptoms of viral infections. However, it should be noted that vaccination releases small amounts of the virus, even if the virus-infected person is asymptomatic or mild. Vaccination does not mean the virus infection is eradicated. Even in COVID-19, if medical doctors do not take measures against infection after carefully analyzing the characteristics of SARS-CoV-2, the prevention policy for the spread of COVID-19 will take the wrong direction.

## Data Availability

Data are available on various websites and have also been made publicly available (more information can be found in the first paragraph of the Results section).

https://kyoto.hosp.go.jp/html/guide/medicalinfo/clinicalresearch/expand/gan.html

## Footnote

All authors have received medical ethics education. In addition, this study has been approved as a clinical medical study at each medical facility. The human serum samples were used in this study, therefore informed consent from the patient is required.

## Ethics Statement

This study was reviewed and approved by the Central Ethics Review Board of the National Hospital Organization of Japan (Meguro, Tokyo, Japan). The approval number for this study is 50-201504. In order to carry out this research, the authors attended a research ethics education course (e-APRIN) conducted by Association for the Promotion of Research Integrity (APRIN; Shinjuku, Tokyo, Japan). The approval numbers of e-APRIN are AP0000151756, AP0000151757, AP0000151758, and AP0000151769.

## Disclosure

The authors declare no potential conflicts of interest. The funders had no role in study design, data collection and analysis, decision to publish, or preparation of the manuscript.

## Acknowledgments

We thank Professor Richard A. Young (Whitehead Institute for Biomedical Research, Massachusetts Institute of Technology, Cambridge, MA, USA) for his assistance with this research. This study was supported in part by grants from the Japan Ministry of Education, Culture, Science, and Technology (Nos. 24592510, 15K1079, and 19K09840); Foundation of Osaka Cancer Research; Ichiro Kanehara Foundation for the Promotion of Medical Sciences and Medical Care; Foundation for Promotion of Cancer Research; Kanzawa Medical Research Foundation; Shinshu Medical Foundation; and Takeda Foundation for Medical Science.

## Author Contributions

T.H. performed most of the experiments and coordinated the project. T.H. and N.Y. conceived the study and wrote the manuscript. N.Y. and I.K. provided expertise in clinical medicine and oversaw the entire study.

## Transparency Document

The transparency document associated with this article can be found in the online version at http://.

## Notes

### Competing Interest Statement

The authors have declared no competing interest.

## References

1. COVID-19: Evolution of the number of cases and deaths. https://interactive.afp.com/graphics/COVID-19-Evolution-du-nombre-de-cas-et-deces_601/

2. Volz E, Mishra S, Chand M, Barrett JC, Johnson R, Geidelberg L, Hinsley WR, Laydon DJ, Dabrera G, O’Toole Á, Amato R, Ragonnet-Cronin M, Harrison I, Jackson B, Ariani CV, Boyd O, Loman NJ, McCrone JT, Gonçalves S, Jorgensen D, Myers R, Hill V, Jackson DK, Gaythorpe K, Groves N, Sillitoe J, Kwiatkowski DP; COVID-19 Genomics UK (COG-UK) consortium, Flaxman S, Ratmann O, Bhatt S, Hopkins S, Gandy A, Rambaut A, Ferguson NM. Assessing transmissibility of SARS-CoV-2 lineage B.1.1.7 in England. Nature. 2021 Mar 25. doi: 10.1038/s41586-021-03470-x.

3. Volz E, Mishra S, Chand M, Barrett JC, Johnson R, Geidelberg L, Hinsley WR, Laydon DJ, Dabrera G, O’Toole Á, Amato R, Ragonnet-Cronin M, Harrison I, Jackson B, Ariani CV, Boyd O, Loman NJ, McCrone JT, Gonçalves S, Jorgensen D, Myers R, Hill V, Jackson DK, Gaythorpe K, Groves N, Sillitoe J, Kwiatkowski DP; COVID-19 Genomics UK (COG-UK) consortium, Flaxman S, Ratmann O, Bhatt S, Hopkins S, Gandy A, Rambaut A, Ferguson NM. Assessing transmissibility of SARS-CoV-2 lineage B.1.1.7 in England. Nature. 2021 Mar 25. doi: 10.1038/s41586-021-03470-x.

4. Firestone MJ, Lorentz AJ, Meyer S, Wang X, Como-Sabetti K, Vetter S, Smith K, Holzbauer S, Beaudoin A, Garfin J, Ehresmann K, Danila R, Lynfield R. First Identified Cases of SARS-CoV-2 Variant P.1 in the United States-Minnesota, January 2021. MMWR Morb Mortal Wkly Rep. 2021 Mar 12;70(10):346–347. doi: 10.15585/mmwr.mm7010e1.

5. Brief report: New Variant Strain of SARS-CoV-2 Identified in Travelers from Brazil. January 12, 2021. https://www.niid.go.jp/niid/en/2019-ncov-e/10108-covid19-33-en.html

6. Emary KRW, Golubchik T, Aley PK, Ariani CV, Angus B, Bibi S, Blane B, Bonsall D, Cicconi P, Charlton S, Clutterbuck EA, Collins AM, Cox T, Darton TC, Dold C, Douglas AD, Duncan CJA, Ewer KJ, Flaxman AL, Faust SN, Ferreira DM, Feng S, Finn A, Folegatti PM, Fuskova M, Galiza E, Goodman AL, Green CM, Green CA, Greenland M, Hallis B, Heath PT, Hay J, Hill HC, Jenkin D, Kerridge S, Lazarus R, Libri V, Lillie PJ, Ludden C, Marchevsky NG, Minassian AM, McGregor AC, Mujadidi YF, Phillips DJ, Plested E, Pollock KM, Robinson H, Smith A, Song R, Snape MD, Sutherland RK, Thomson EC, Toshner M, Turner DPJ, Vekemans J, Villafana TL, Williams CJ, Hill AVS, Lambe T, Gilbert SC, Voysey M, Ramasamy MN, Pollard AJ; COVID-19 Genomics UK consortium; AMPHEUS Project; Oxford COVID-19 Vaccine Trial Group. Efficacy of ChAdOx1 nCoV-19 (AZD1222) vaccine against SARS-CoV-2 variant of concern 202012/01 (B.1.1.7): an exploratory analysis of a randomised controlled trial. Lancet. 2021 Mar 30:S0140-6736(21)00628-0. doi: 10.1016/S0140-6736(21)00628-0.

7. Planas D, Bruel T, Grzelak L, Guivel-Benhassine F, Staropoli I, Porrot F, Planchais C, Buchrieser J, Rajah MM, Bishop E, Albert M, Donati F, Prot M, Behillil S, Enouf V, Maquart M, Smati-Lafarge M, Varon E, Schortgen F, Yahyaoui L, Gonzalez M, De Sèze J, Péré H, Veyer D, Sève A, Simon-Lorière E, Fafi-Kremer S, Stefic K, Mouquet H, Hocqueloux L, van der Werf S, Prazuck T, Schwartz O. Sensitivity of infectious SARS-CoV-2 B.1.1.7 and B.1.351 variants to neutralizing antibodies. Nat Med. 2021 Mar 26. doi: 10.1038/s41591-021-01318-5.

8. Hoffmann M, Arora P, Groß R, Seidel A, Hörnich BF, Hahn AS, Krüger N, Graichen L, Hofmann-Winkler H, Kempf A, Winkler MS, Schulz S, Jäck HM, Jahrsdörfer B, Schrezenmeier H, Müller M, Kleger A, Münch J, Pöhlmann S. SARS-CoV-2 variants B.1.351 and P.1 escape from neutralizing antibodies. Cell. 2021 Mar 20:S0092-8674(21)00367-6. doi: 10.1016/j.cell.2021.03.036.

9. COVID-19 – Global. Posted on WHO website Posted on 31 December 2021. https://www.who.int/emergencies/disease-outbreak-news/item/2020-DON305

10. Rikhtegaran Tehrani Z, Saadat S, Saleh E, et al. Performance of nucleocapsid and spike-based SARS-CoV-2 serologic assays. PLoS One. 2020;15(11):e0237828. doi:10.1371/journal.pone.0237828

11. Yuan M. Liu H. Wu NC. Lee CD. Zhu X. Zhao F. Huang D. Yu W. Hua Y. Tien H. Rogers TF. Landais E. Sok D. Jardine JG. Burton DR. Wilson IA. Structural basis of a shared antibody response to SARS-CoV-2. Science 369 (6507): 1119-1123 (2020)

12. Weinreich DM, Sivapalasingam S, Norton T, Ali S, Gao H, Bhore R, Musser BJ, Soo Y, Rofail D, Im J, Perry C, Pan C, Hosain R, Mahmood A, Davis JD, Turner KC, Hooper AT, Hamilton JD, Baum A, Kyratsous CA, Kim Y, Cook A, Kampman W, Kohli A, Sachdeva Y, Graber X, Kowal B, DiCioccio T, Stahl N, Lipsich L, Braunstein N, Herman G, Yancopoulos GD; Trial Investigators. REGN-COV2, a Neutralizing Antibody Cocktail, in Outpatients with Covid-19. N Engl J Med. 2021 Jan 21;384(3):238–251. doi: 10.1056/NEJMoa2035002

13. Takahashi T, Ellingson MK, Wong P, Israelow B, Lucas C, Klein J, Silva J, Mao T, Oh JE, Tokuyama M, Lu P, Venkataraman A, Park A, Liu F, Meir A, Sun J, Wang EY, Casanovas-Massana A, Wyllie AL, Vogels CBF, Earnest R, Lapidus S, Ott IM, Moore AJ; Yale IMPACT Research Team, Shaw A, Fournier JB, Odio CD, Farhadian S, Dela Cruz C, Grubaugh ND, Schulz WL, Ring AM, Ko AI, Omer SB, Iwasaki A. Sex differences in immune responses that underlie COVID-19 disease outcomes. Nature 2020 Dec;588(7837):315–320. doi: 10.1038/s41586-020-2700-3.

14. Yang F, Nielsen SCA, Hoh RA, Röltgen K, Wirz OF, Haraguchi E, Jean GH, Lee JY, Pham TD, Jackson KJL, Roskin KM, Liu Y, Nguyen K, Ohgami RS, Osborne EM, Nadeau KC, Niemann CU, Parsonnet J, Boyd SD. Shared B cell memory to coronaviruses and other pathogens varies in human age groups and tissues.Science.2021 Apr 12:eabf6648. doi: 10.1126/science.abf6648.

15. Chen RE, Zhang X, Case JB, Winkler ES, Liu Y, VanBlargan LA, Liu J, Errico JM, Xie X, Suryadevara N, Gilchuk P, Zost SJ, Tahan S, Droit L, Turner JS, Kim W, Schmitz AJ, Thapa M, Wang D, Boon ACM, Presti RM, O’Halloran JA, Kim AHJ, Deepak P, Pinto D, Fremont DH, Crowe JE Jr, Corti D, Virgin HW, Ellebedy AH, Shi PY, Diamond MS. Resistance of SARS-CoV-2 variants to neutralization by monoclonal and serum-derived polyclonal antibodies. Nat Med. 2021 Apr;27(4):717–726. doi: 10.1038/s41591-021-01294-w.

16. Tegally H, Wilkinson E, Giovanetti M, Iranzadeh A, Fonseca V, Giandhari J, Doolabh D, Pillay S, San EJ, Msomi N, Mlisana K, von Gottberg A, Walaza S, Allam M, Ismail A, Mohale T, Glass AJ, Engelbrecht S, Van Zyl G, Preiser W, Petruccione F, Sigal A, Hardie D, Marais G, Hsiao NY, Korsman S, Davies MA, Tyers L, Mudau I, York D, Maslo C, Goedhals D, Abrahams S, Laguda-Akingba O, Alisoltani-Dehkordi A, Godzik A, Wibmer CK, Sewell BT, Lourenço J, Alcantara LCJ, Kosakovsky Pond SL, Weaver S, Martin D, Lessells RJ, Bhiman JN, Williamson C, de Oliveira T. Detection of a SARS-CoV-2 variant of concern in South Africa. Nature. 2021 Mar 9. doi: 10.1038/s41586-021-03402-9.

17. Maggi F, Novazzi F, Genoni A, Baj A, Spezia PG, Focosi D, Zago C, Colombo A, Cassani G, Pasciuta R, Tamborini A, Rossi A, Prestia M, Capuano R, Azzi L, Donadini A, Catanoso G, Grossi PA, Maffioli L, Bonelli G. Imported SARS-CoV-2 Variant P.1 in Traveler Returning from Brazil to Italy. Emerg Infect Dis. 2021 Apr;27(4):1249–1251. doi: 10.3201/eid2704.210183.

18. SARS-CoV-2 Variant – United Kingdom of Great Britain and Northern Ireland. Emergencies preparedness, response. WHO Disease Outbreak News. 21 December 2020. https://www.who.int/csr/don/21-december-2020-sars-cov2-variant-united-kingdom/en

19. Hayashi T, Konishi I. Research Letter: Spread of Infection With Tokyo-type SARS-CoV-2 Variant (E484K). JAMA. Published on online April 07, 2021. https://jamanetwork.com/journals/jama/fullarticle/2776543

20. Hayashi T. Konishi I. In Tokyo, the situation of the spread of SARS-CoV-2 variants. Science. Vol. 372, Issue 6538. pp. 1–2. Published on April 08, 2021. https://science.sciencemag.org/content/372/6538/eabg3055/tab-e-letters

