## supplemental material 1 for "Neutralizing response against E484K variant after original SARS-CoV-2 infection"

### Effect of RBD mutations in spike glycoprotein of SARS-CoV-2 on neutralizing IgG affinity

#### Materials and Methods

##### Adsorption of conventional original RBD, RBD Y453F, RBD N501Y and RBD E484K recombinant proteins to the solid phase surface of the ELISA plate

HeLa cells were transfected with either pcMV3-2019-nCov-RBD-Flag tag expression vector (2 mg), pcMV3-2019-nCov-RBD Y453F-Flag tag expression vector (2 mg), or pcMV3-2019-nCov-RBD N501Y-Flag tag expression vector (2 mg) (Sino Biological Inc. Beijing, China). The cells transfected with 2019-nCov-RBD, 2019-nCov-RBD Y453F, or 2019-nCov-RBD N501Y expression vectors were incubated for 48 h prior to harvesting. The conventional RBD, Y453F, and N501Y recombinant proteins were purified using a His-tag column following the standard procedure. Purified conventional RBD, Y453F, or N501Y recombinant proteins were adsorbed on the solid phase surface of the ELISA plate (SMILON ELISA plate MS-3508F, Tokyo Japan).

##### Quantitative measurement of neutralizing antibodies to original SARS-CoV-2 or SARS-CoV-2 variants

The SeroFlash™ SARS-CoV-2 Neutralizing Antibody Assay Fast Kit (EpiGentek Group Inc., NY) contains all the reagents necessary for quantitatively measuring the level of SARS-CoV-2 neutralizing antibodies. In this assay, conventional RBD, Y453F, or N501Y spike proteins were stably pre-coated onto microplate wells. His-tagged ACE2 binds to the coated spike protein in the presence or absence of neutralizing antibodies in the sample. The amount of the bound ACE2, which is proportional to ACE2 inhibition intensity, is then recognized by the neutralizing detection complex, which contains anti-His antibodies and is measured through an ELISA-like reaction, where the absorbance is read by a microplate spectrophotometer at a wavelength of 450 nm. The neutralizing antibody level is inversely proportional to the optical density intensity measured, i.e., the higher the level of neutralizing antibody, the lower the OD intensity. Quantitative measurements of SARS-CoV-2 neutralizing antibody levels were performed according to the manufacturer's procedure.

**Qualitative measurement of SARS-CoV-2 neutralizing antibodies**

Serum samples (25 µL) from 20 COVID-19-negative patients and 41 COVID-19-positive patients with varying immunoglobulin M (IgM) and immunoglobulin G (IgG) antibody levels were used for this assay. COVID-19 status was confirmed by RT-PCR, antigen, and/or antibody serology tests. All serum samples were provided by RayBiotech (RayBiotech Life, GA). The details of the materials and methods used are described in the supplementary materials. A neutralizing inhibition score of ≥ 20% was used to indicate a positive result and detection of SARS-CoV-2 neutralizing antibody; a score under 20% was used to indicate a negative result and no detectable SARS-CoV-2 neutralizing antibody.

#### Materials and Methods

##### Analyzes the three-dimensional structure of the binding site between mink and human ACE2

**Spanner** is a structural homology modeling pipeline that threads a query amino-acid sequence onto a template protein structure. Spanner is unique in that it handles gaps by spanning the region of interest using fragments of known structures.
To create a model, you must provide a template structure, as well as an alignment of the query sequence you wish to model onto the template sequence. Spanner will replace mismatched residues, and fill any gaps caused by insertions or deletions.

For users that are unable to create an alignment a method for building a model starting only from sequence is also available. During this process a template search is conducted and an alignment is built dynamically using FORTE before being passed through to the main part of the pipeline.

Spanner consists of several modules written in the Go programming language. For Spanner jobs which build a model only from sequence, the first step is a search of the PDB for possible templates using BLAST. These possible templates are then aligned and scored with FORTE.

The next step involves defining the start and end points of fragments corresponding to insertions or deletions. The start and end points are referred to as anchors because they must be equivalent in both the template and any candidate fragment. The margin parameter determines how far from the edge of a gap the fragment begins or ends. For example a margin of 0 would mean that the anchors begins at the very edge of a gap. This is usually not a good idea, and the default margin is set to 1.
A representative set of protein chains was prepared using CD-HIT at 100% sequence identity.[3] All continuous fragments were then extracted from this set of chains and stored in a relational database, indexed by the internal coordinates of the fragment endpoints. A separate database is prepared for each fragment length. Currently, fragments of length 8-40, including the 8 anchor residues, are stored in the database.

**Cn3D** ("see in 3D") is a helper application for your web browser that allows researcher to view 3-dimensional structures from NCBI's Entrez Structure database. Cn3D is provided for Windows and Macintosh, and can be compiled on Unix. Cn3D simultaneously displays structure, sequence, and alignment, and now has powerful annotation and alignment editing features. *(For those who prefer to view 3D structures on the web, without the need to install a separate application, iCn3D ("I see in 3D") is available as of April 2016.)*

•**Web-based Structure Viewer**iCn3D ("I see in 3D"), released in April 2016, provides interactive views of three-dimensional macromolecular structures on the web.

•There is no need to install a separate application in order to use iCn3D; you just need to use a web browser that supports WebGL.

•iCn3D also allows you to cutomize the display of a structure and generate a URL that allows you to share the link, and to incorporate iCn3D into your own web pages.

•**New Features in Cn3D 4.3.1:**View superpositions of structures that have similar molecular complexes, as identified by the newly released VAST+ (an enhanced version of the existing Vector Alignment Search Tool). The VAST+ help document provides additional details about the tool and examples of how it can be used to learn more about proteins.

•Cn3D 4.3.1 uses the MIME type: application/vnd.ncbi.cn3d.

Analyses of protein contact residues and protein buried surface areas Protein contact residues were analyzed using the **LigPlot+ program (v.1.4.5)** (https://www.ebi.ac.uk/thornton-srv/software/ LigPlus/). Protein buried surface areas were analysed using **PDBePISA tool** (http://pdbe.org/pisa/) and **MOE project DB for Protein-Protein Docking** (MOLSIS Inc. Tokyo Japan). The modeling and Docking of the mink ACE2 protein and RBD in Spike Glycoprotein SARS-CoV-2 was analyzed by **MOE project DB** with previously posted ID PDB and protein ID (MOLSIS Inc. Tokyo Japan). The binding affinity between mink ACE2 and RDB in Spike Glycoprotein of SRARS-CoV-2 was analyzed by **MOE project DB** (MOLSIS Inc.).

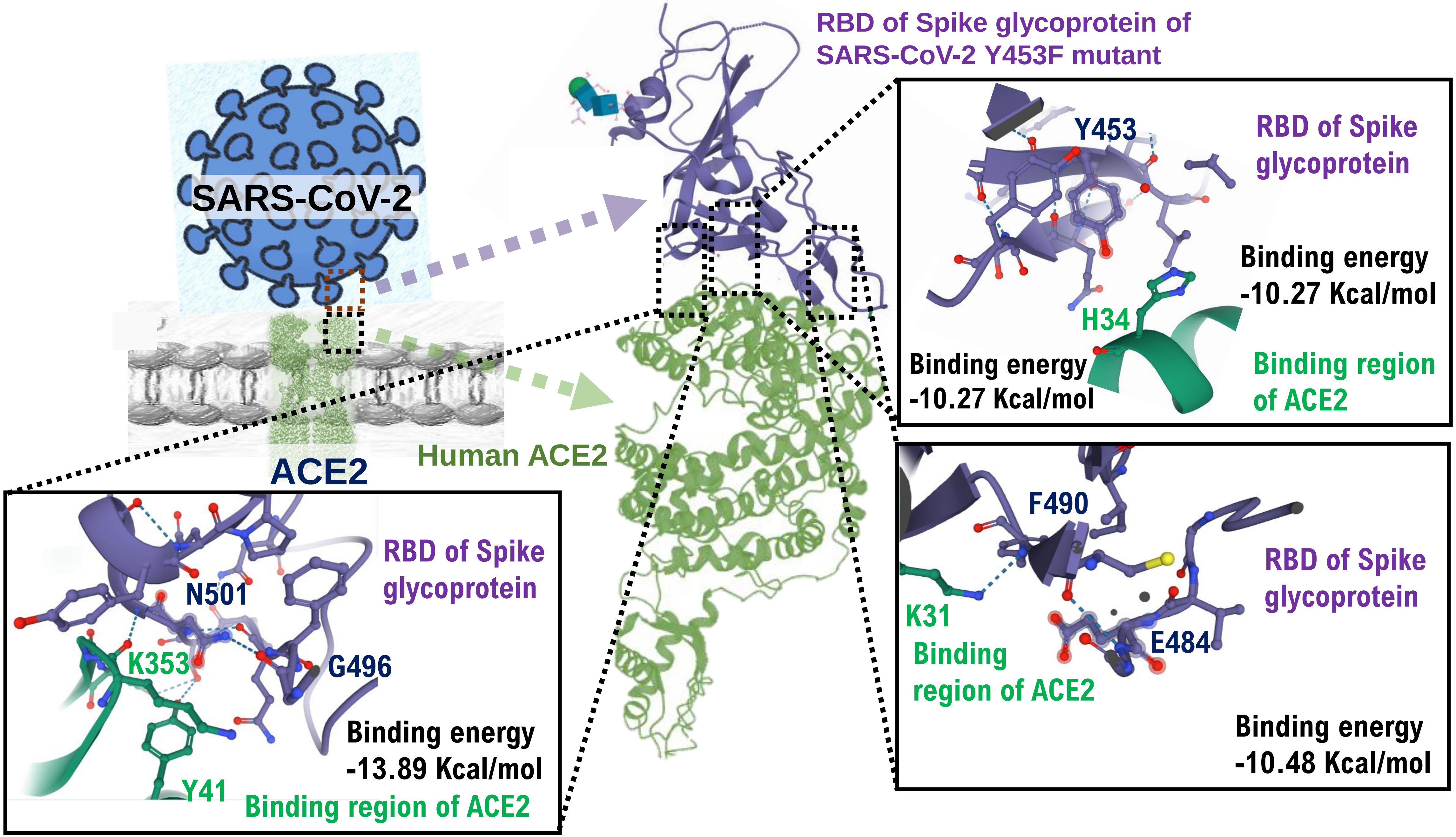
RBD of Spike glycoprotein of SARS-CoV-2 Y453F mutant
SARS-CoV-2
ACE2
Human ACE2
Y453
RBD of Spike glycoprotein
Binding energy -10.27 Kcal/mol
H34
Binding energy -10.27 Kcal/mol
Binding region of ACE2
N501
RBD of Spike glycoprotein
K353
G496
Binding energy -13.89 Kcal/mol
Y41
Binding region of ACE2
F490
RBD of Spike glycoprotein
K31
Binding region of ACE2
E484
Binding energy -10.48 Kcal/mol

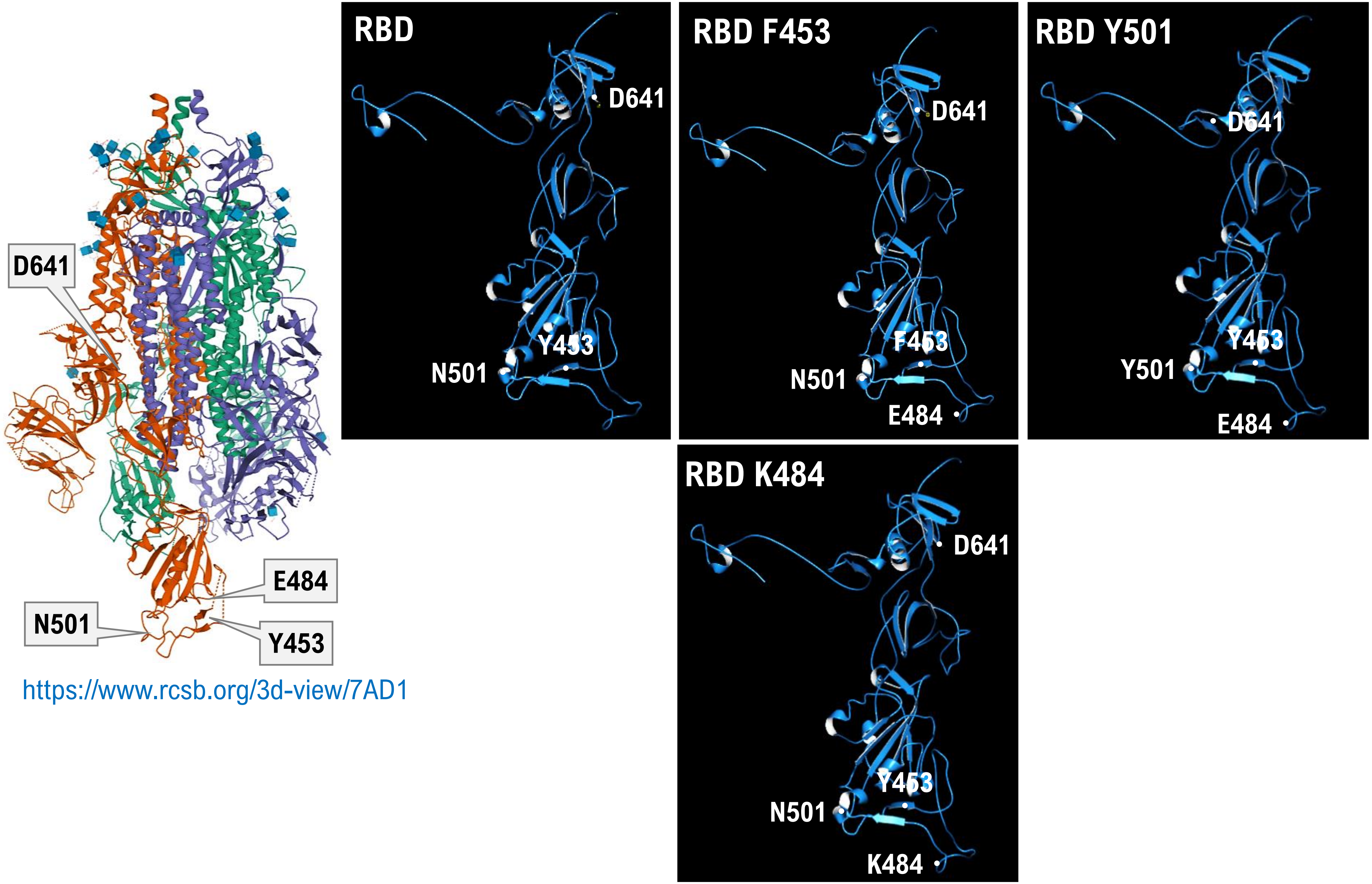


https://www.rcsb.org/3d-view/7AD1

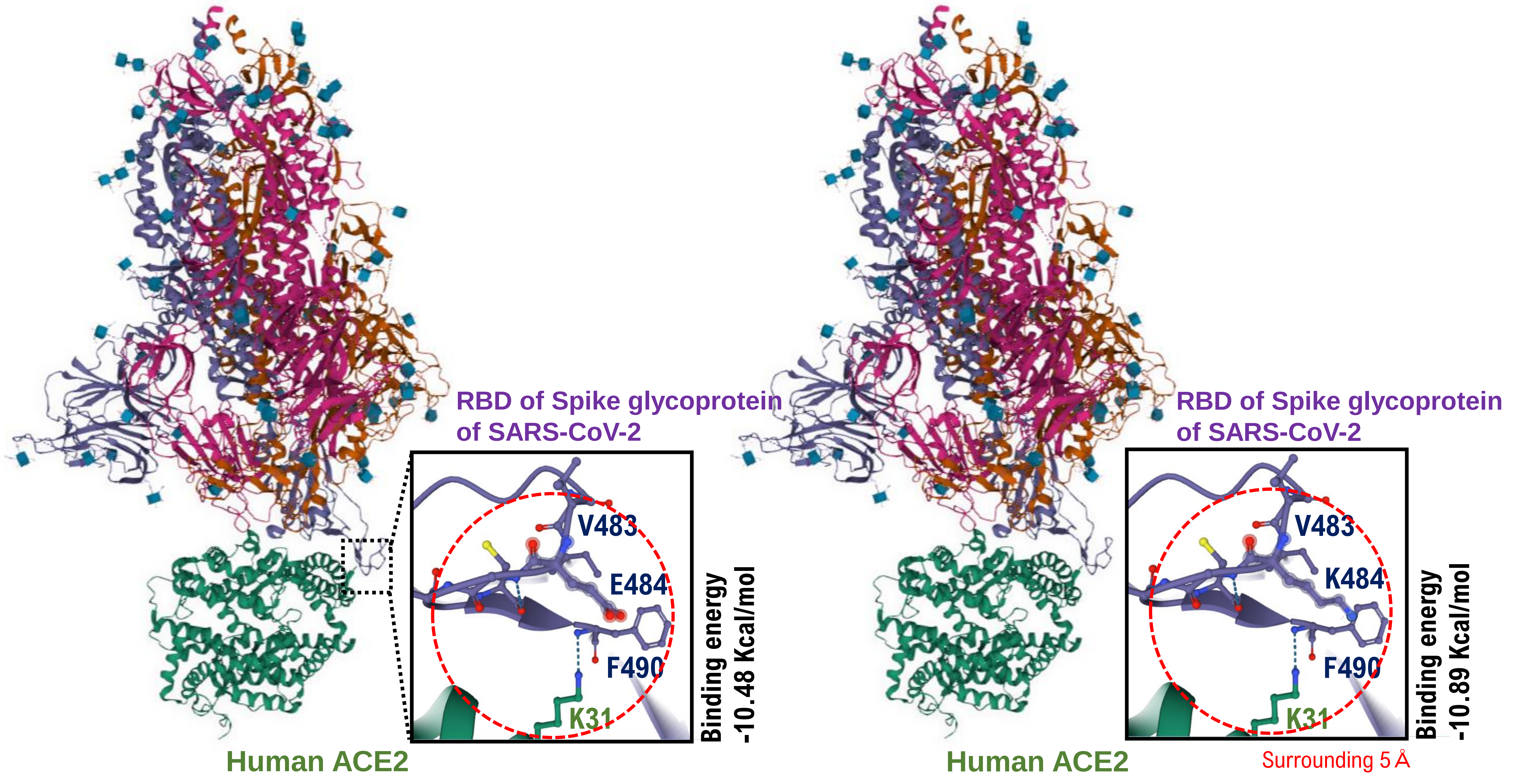
RBD of Spike glycoprotein
of SARS-CoV-2
V483
E484
F490
K31
Binding energy
-10.48 Kcal/mol
Human ACE2
RBD of Spike glycoprotein
of SARS-CoV-2
V483
K484
F490
K31
Binding energy
-10.89 Kcal/mol
Human ACE2
Surrounding 5 Å

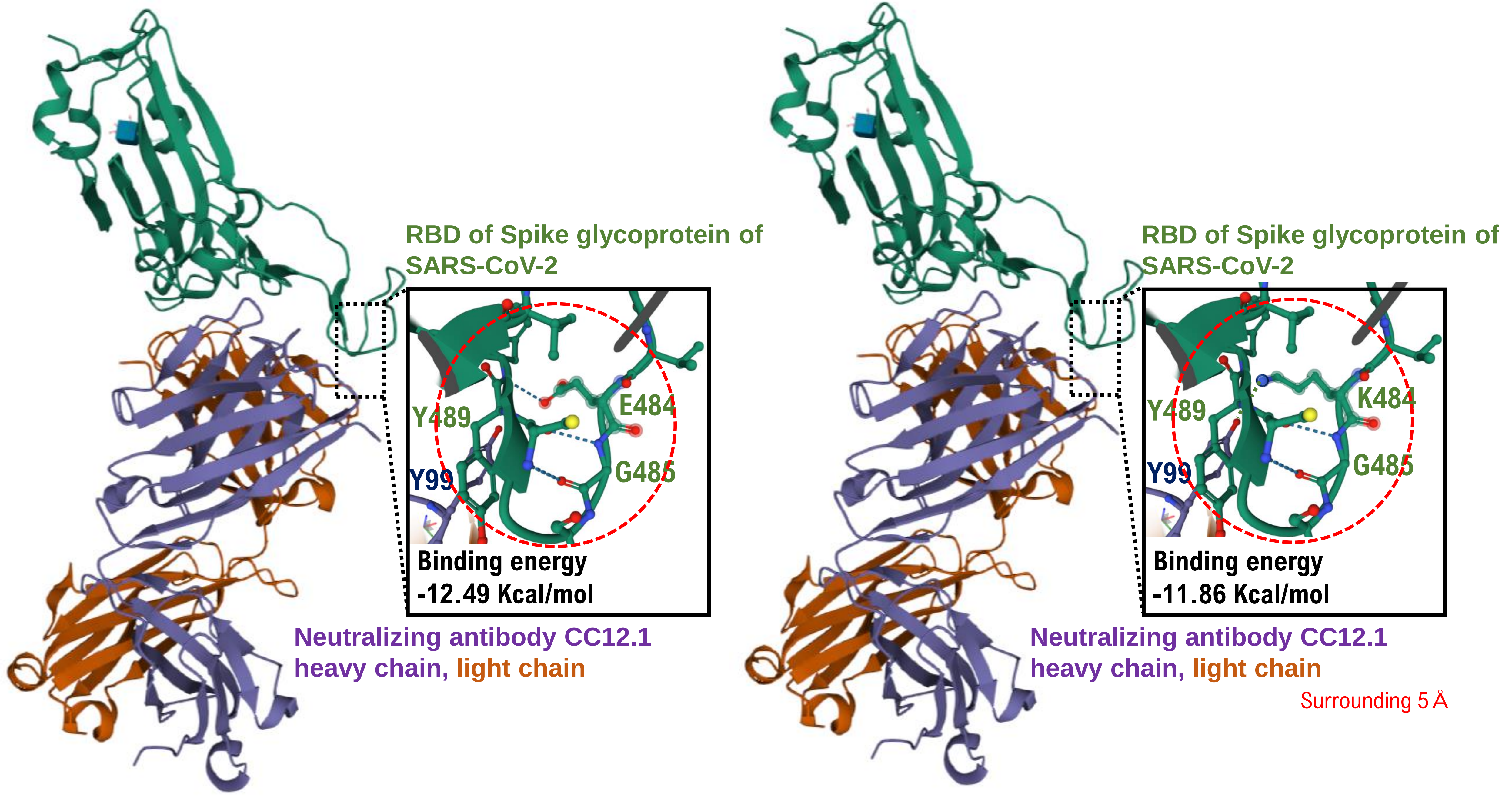
RBD of Spike glycoprotein of SARS-CoV-2
Y489
E484
Y99
G485
Binding energy -12.49 Kcal/mol
Neutralizing antibody CC12.1
heavy chain, light chain
RBD of Spike glycoprotein of SARS-CoV-2
Y489
K484
Y99
G485
Binding energy -11.86 Kcal/mol
Neutralizing antibody CC12.1
heavy chain, light chain
Surrounding 5 Å

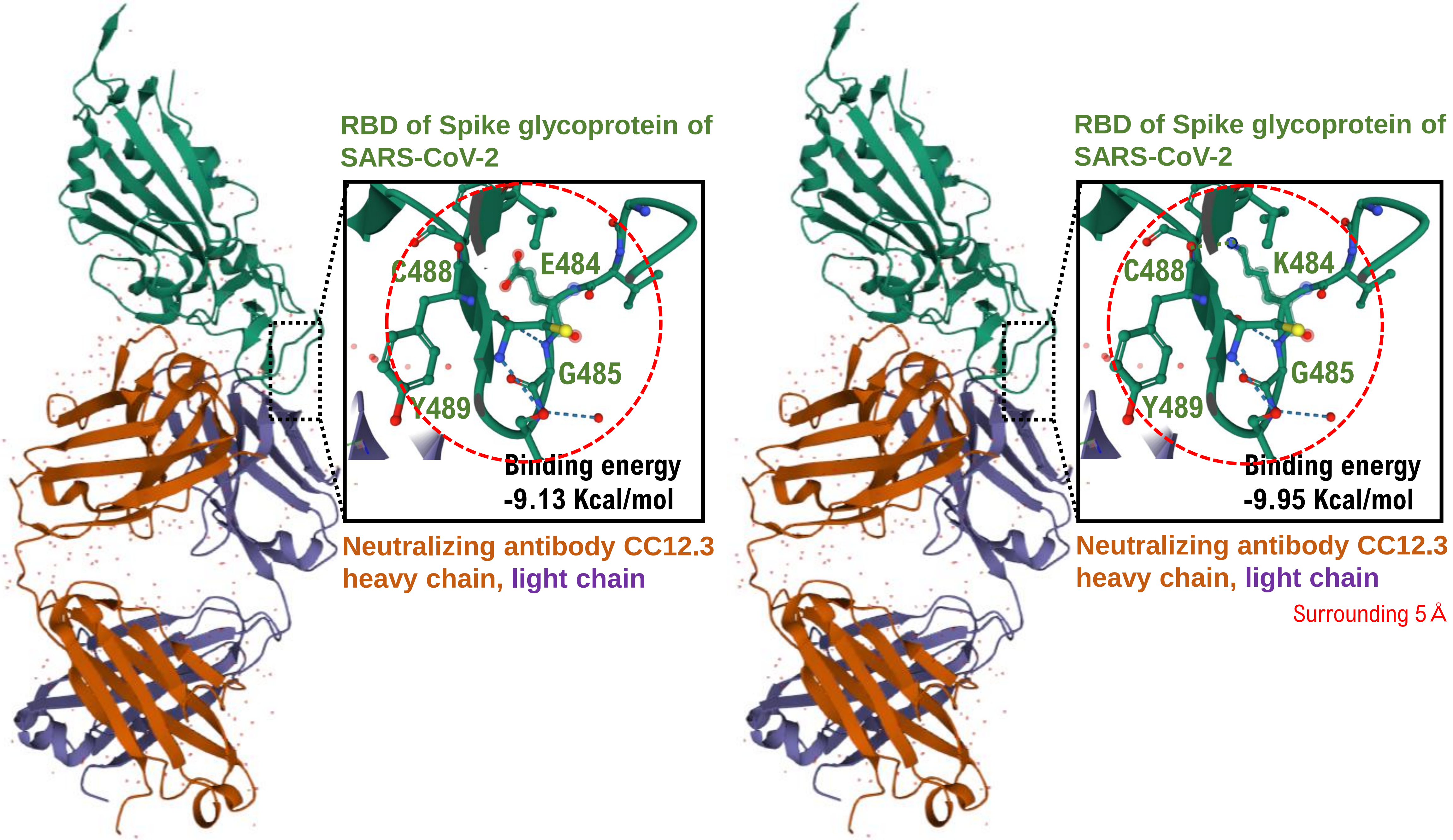
RBD of Spike glycoprotein of SARS-CoV-2
C488
E484
G485
Y489
Binding energy -9.13 Kcal/mol
Neutralizing antibody CC12.3 heavy chain, light chain
RBD of Spike glycoprotein of SARS-CoV-2
C488
K484
G485
Y489
Binding energy -9.95 Kcal/mol
Neutralizing antibody CC12.3 heavy chain, light chain
Surrounding 5 Å

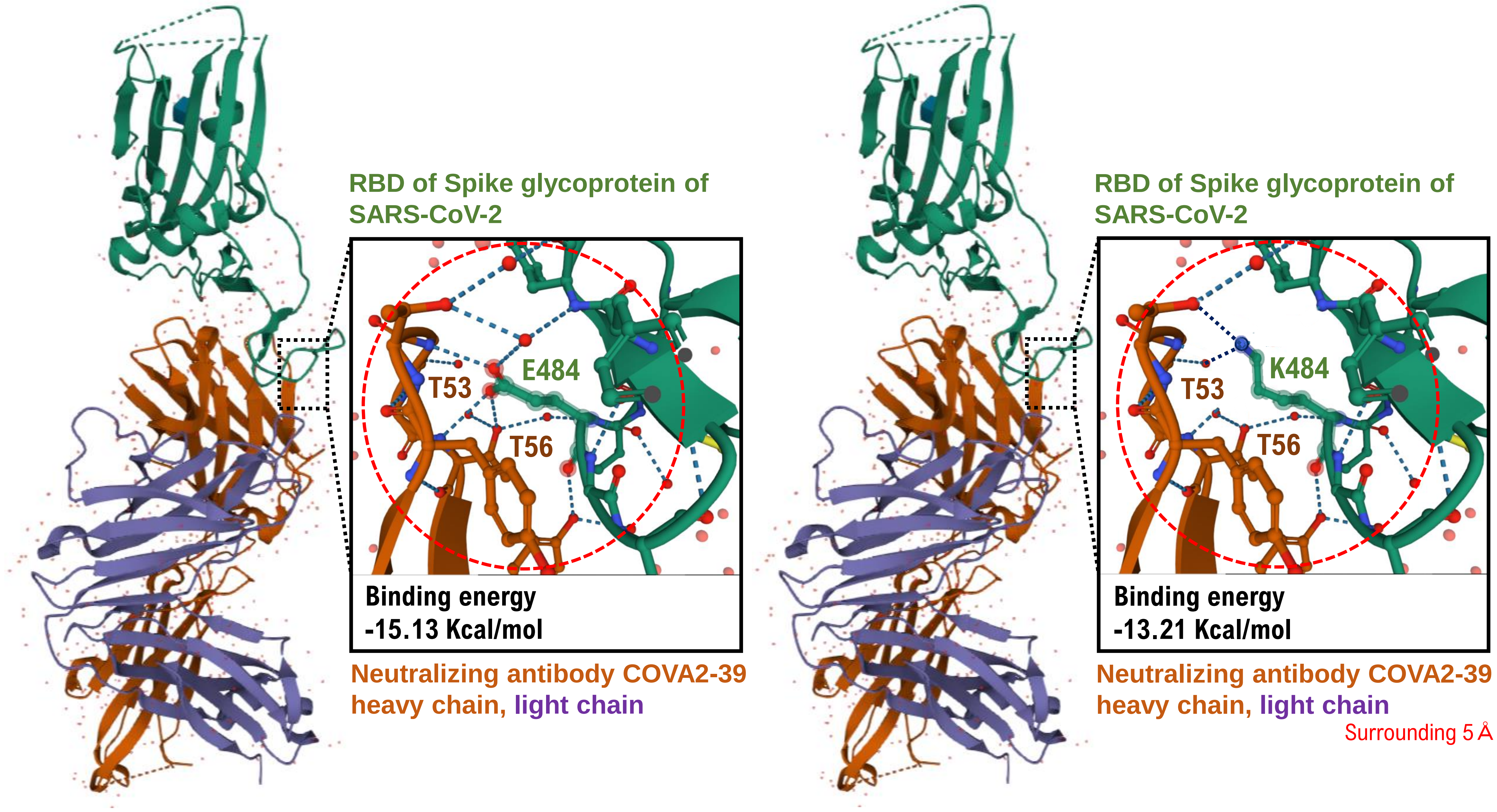
RBD of Spike glycoprotein of SARS-CoV-2
E484
T53
T56
Binding energy
-15.13 Kcal/mol
Neutralizing antibody COVA2-39 heavy chain, light chain
RBD of Spike glycoprotein of SARS-CoV-2
K484
T53
T56
Binding energy
-13.21 Kcal/mol
Neutralizing antibody COVA2-39 heavy chain, light chain
Surrounding 5 Å

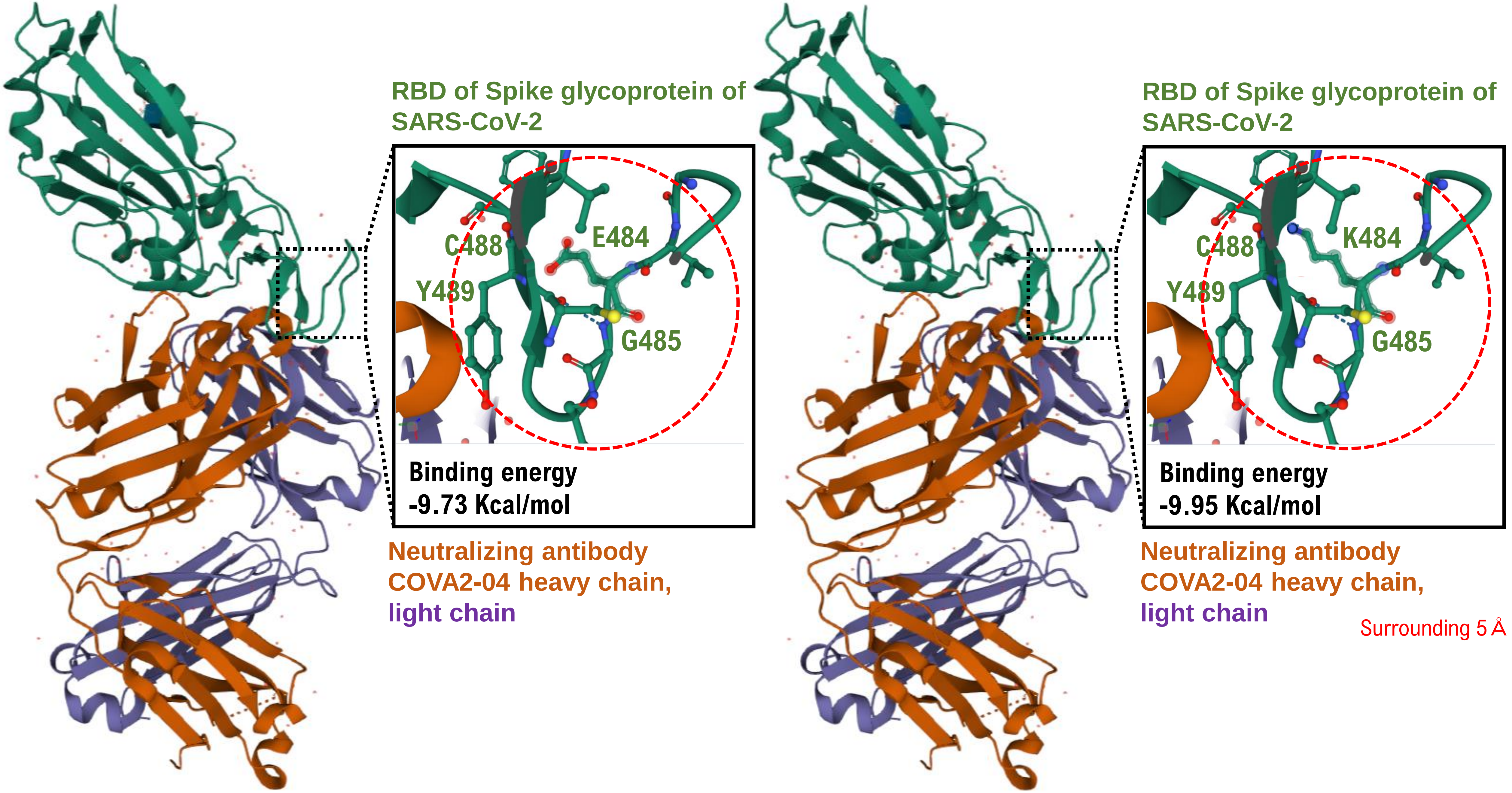
RBD of Spike glycoprotein of SARS-CoV-2
C488
E484
Y489
G485
Binding energy -9.73 Kcal/mol
Neutralizing antibody COVA2-04 heavy chain, light chain
RBD of Spike glycoprotein of SARS-CoV-2
C488
K484
Y489
G485
Binding energy -9.95 Kcal/mol
Neutralizing antibody COVA2-04 heavy chain, light chain
Surrounding 5 Å

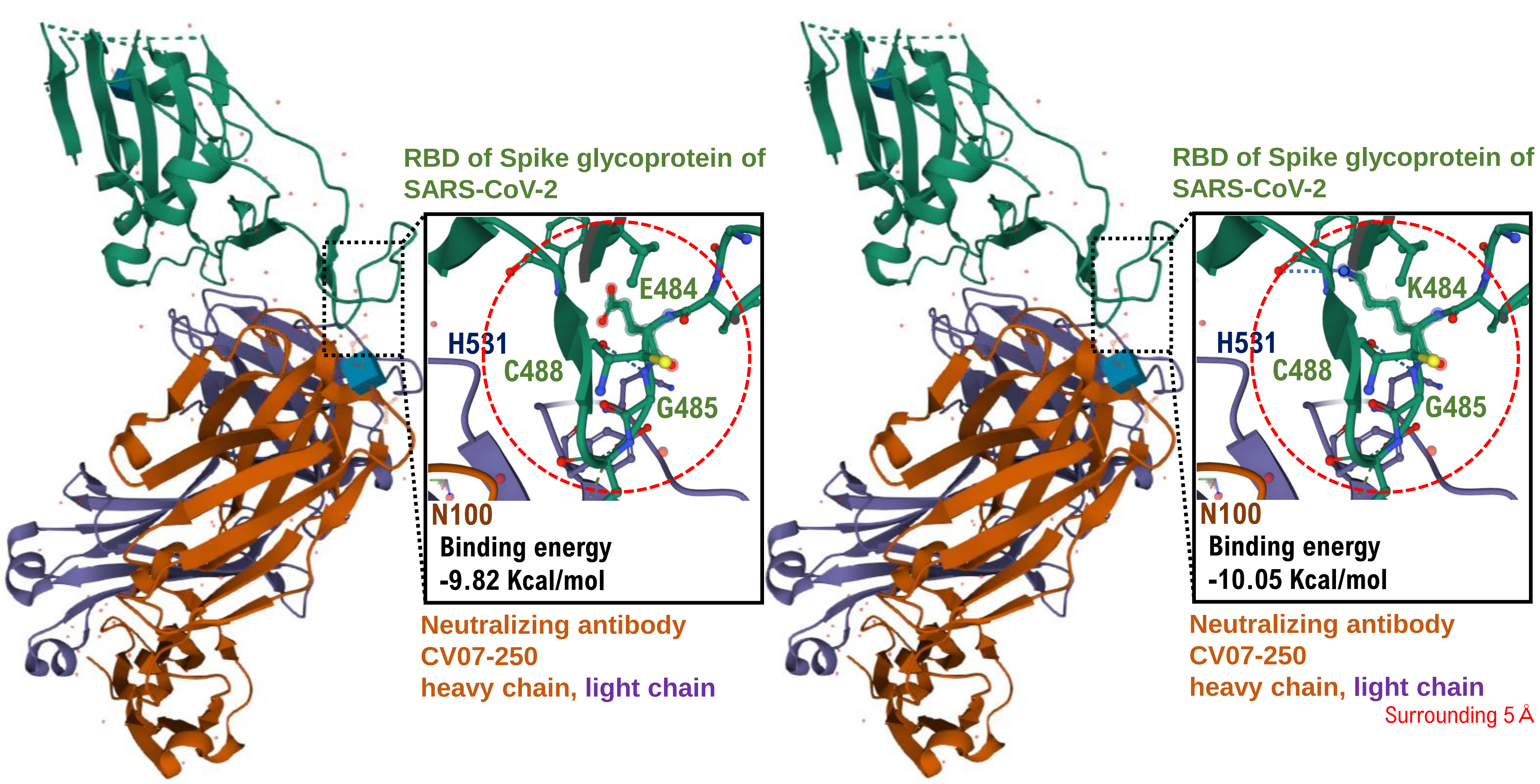
RBD of Spike glycoprotein of SARS-CoV-2
E484
H531
C488
G485
N100
Binding energy
-9.82 Kcal/mol
Neutralizing antibody
CV07-250
heavy chain, light chain
RBD of Spike glycoprotein of SARS-CoV-2
K484
H531
C488
G485
N100
Binding energy
-10.05 Kcal/mol
Neutralizing antibody
CV07-250
heavy chain, light chain
Surrounding 5 Å

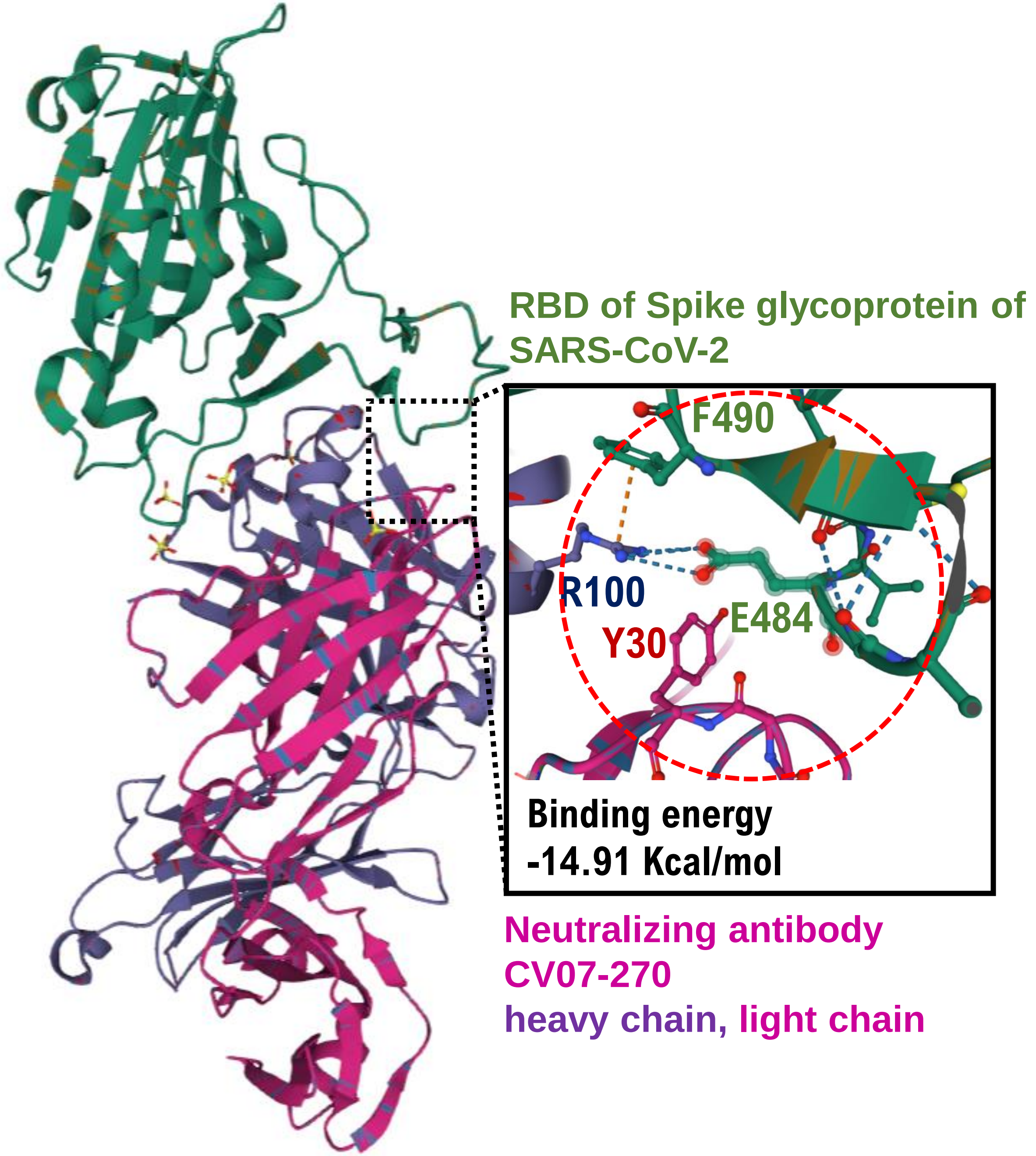
RBD of Spike glycoprotein of SARS-CoV-2
F490
R100
E484
Y30
Binding energy
-14.91 Kcal/mol
Neutralizing antibody
CV07-270
heavy chain, light chain

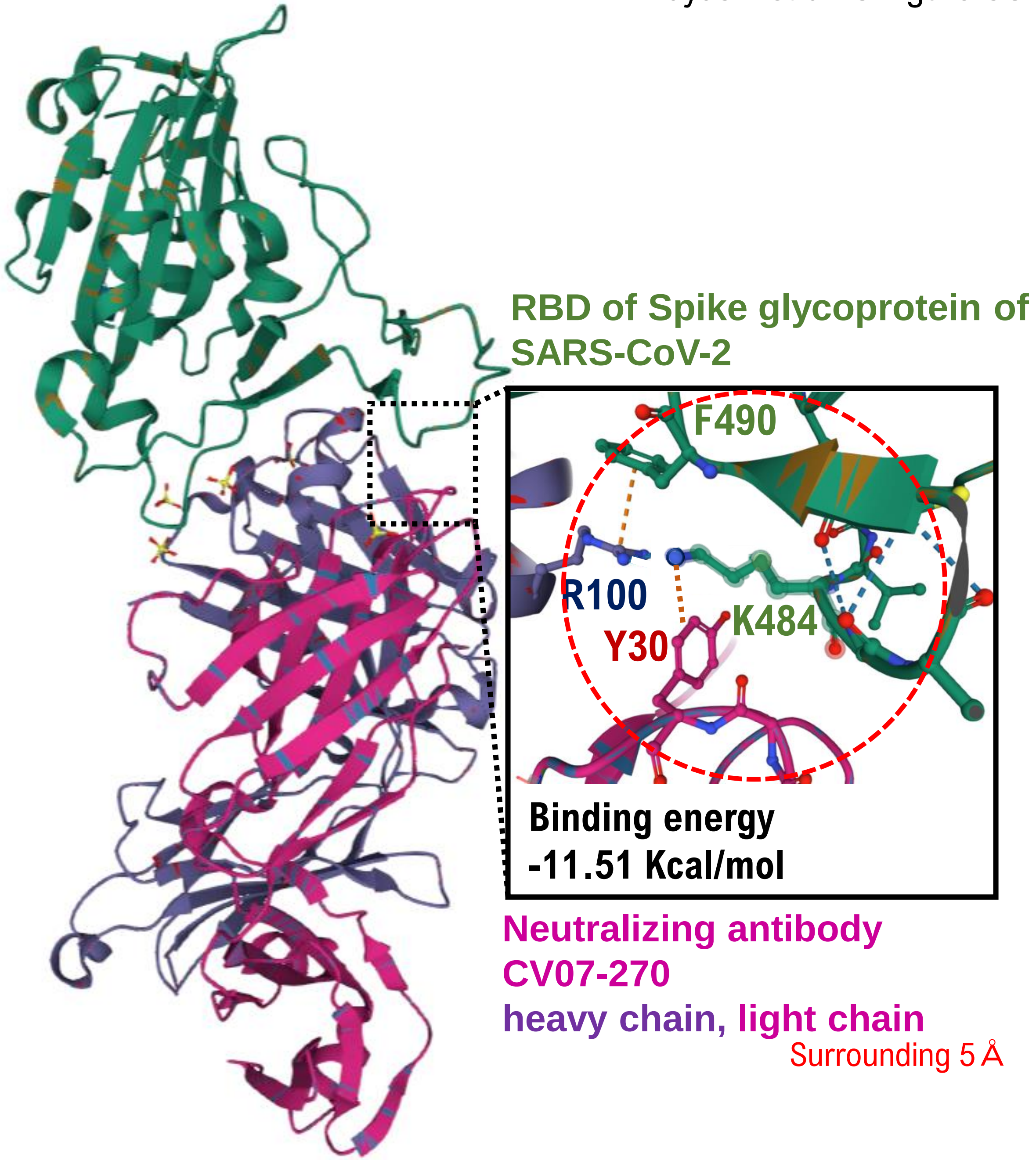
RBD of Spike glycoprotein of SARS-CoV-2
F490
R100
K484
Y30
Binding energy
-11.51 Kcal/mol
Neutralizing antibody
CV07-270
heavy chain, light chain
Surrounding 5 Å

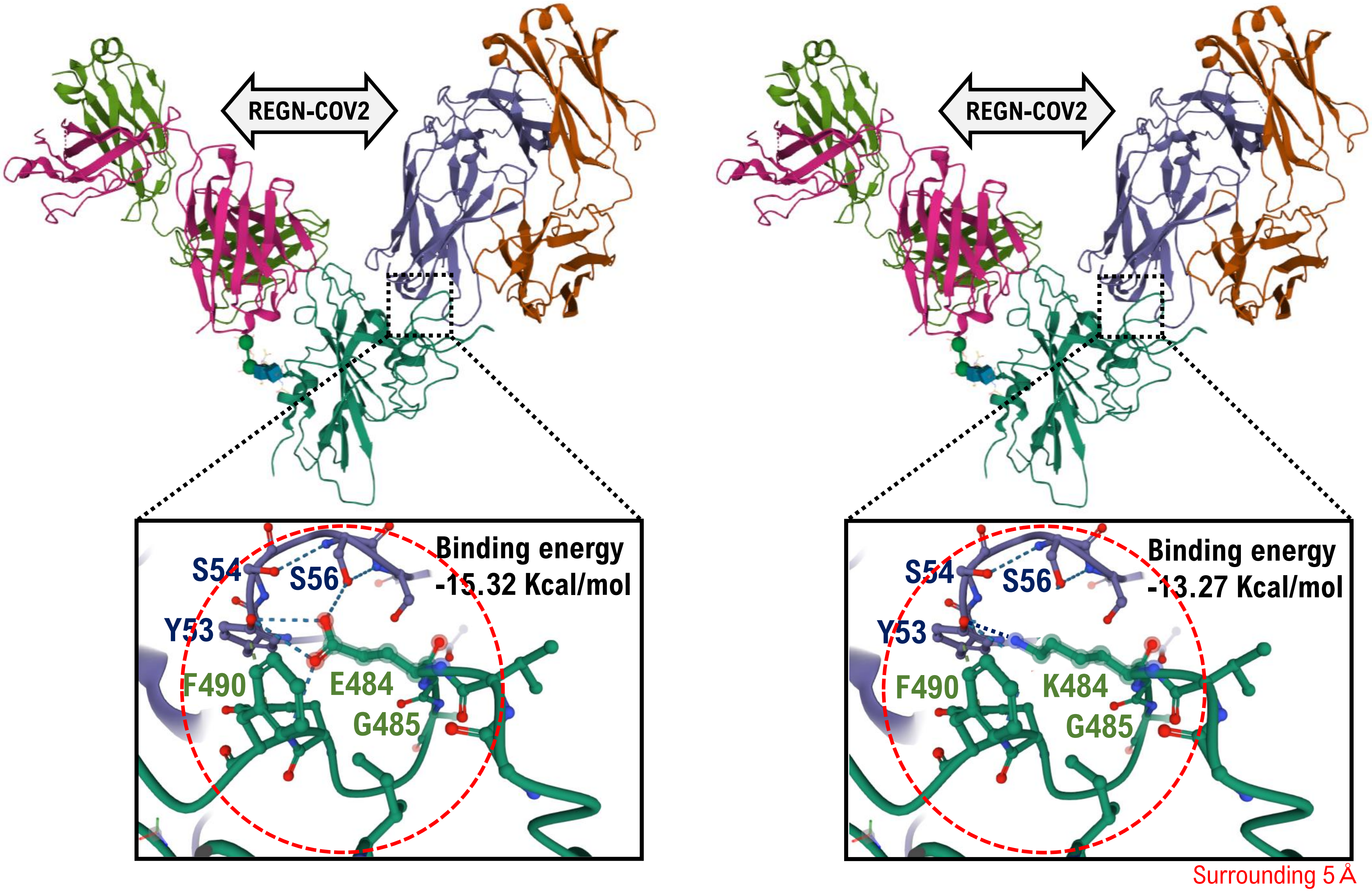
REGN-COV2
S54
S56
Binding energy
-15.32 Kcal/mol
Y53
F490
E484
G485
REGN-COV2
S54
S56
Binding energy
-13.27 Kcal/mol
Y53
F490
K484
G485
Surrounding 5 Å

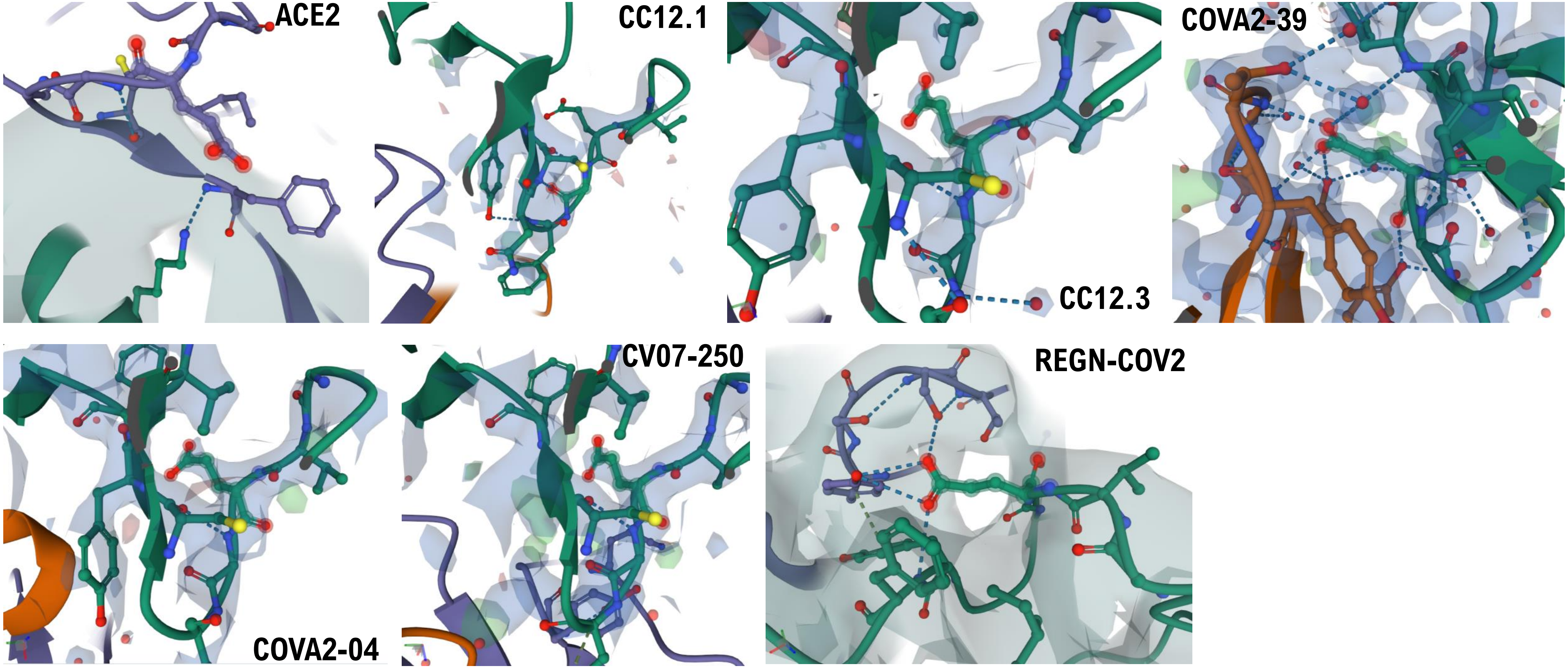


**Electron density map.** 2*F*o−*F*c electron density maps contoured at 1.5$\sigma$ at the binding interface between the SARS-CoV-2 RBD and human ACE2 or the SARS-CoV-2 RBD and neutoralizing antibodies.

Hayashi *et al*. S.Figure 4A

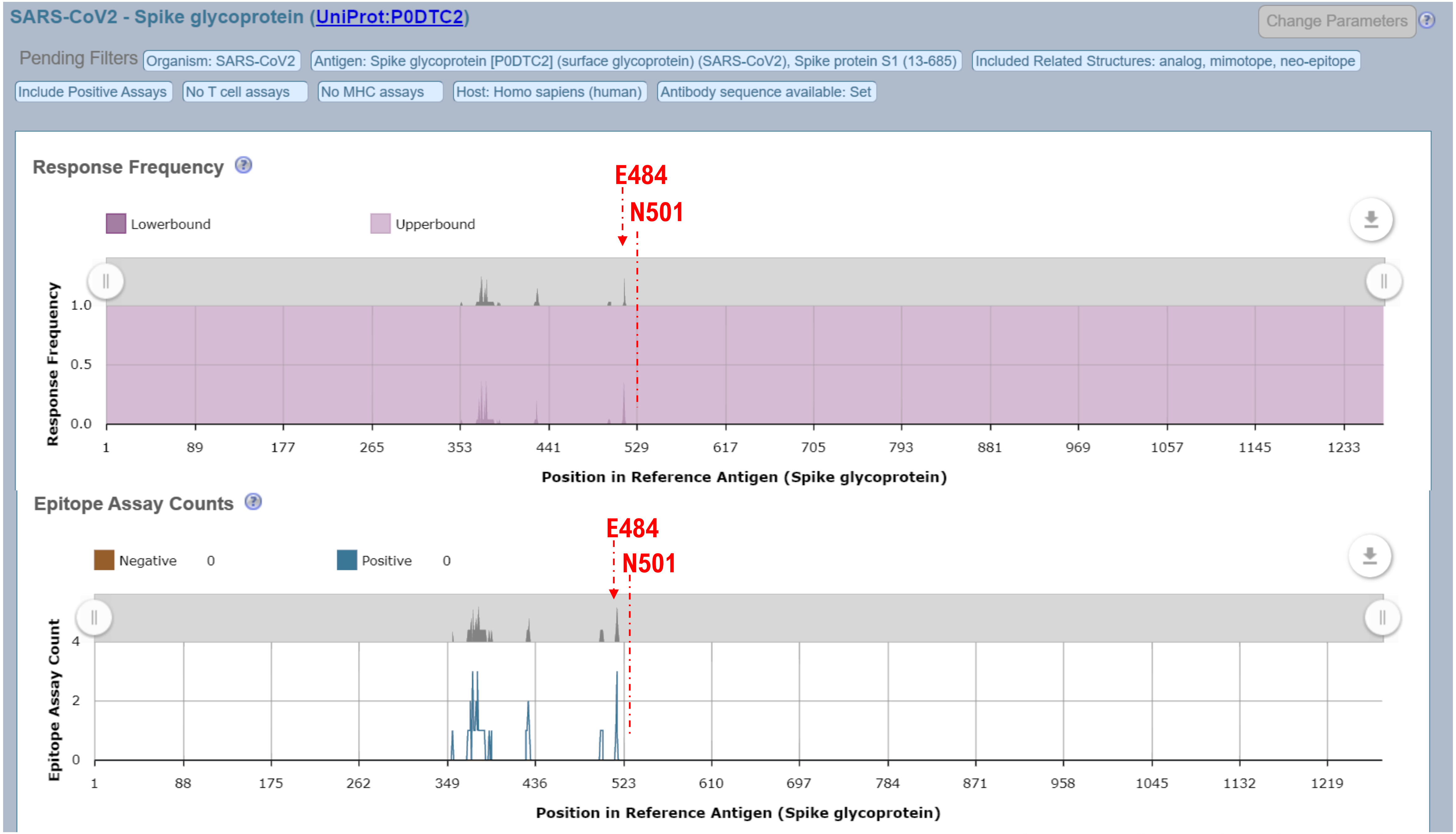


http://www.iedb.org/result_v3.php?cookie_id=6e2edb

Results Returned: 6 | Displaying: 6 Display Graphed Residue Positions Export Results

| Epitope ID | Epitope Sequence | Mapped Position | Identity | Subjects Tested | Subjects Responded | Assays Positive | Assays Negative | Response Freq. (95% CI) |
|---|---|---|---|---|---|---|---|---|
| 1087821 | N354 | 354-354 | 100% | 1 | 1 | 1 | 0 | 1.00 (0.04:1.00) |
| 997006 | Y369, N370, S371, A372, F374, F377, K378, C379, Y380, G381, V382, S383, P384, T385, K386, L390, F429, T430, F515, E516, L517 | 369-517 | 100% | 1 | 1 | 1 | 0 | 1.00 (0.04:1.00) |
| 1125015 | A372, F374, C379 | 372-379 | 100% | 1 | 1 | 1 | 0 | 1.00 (0.04:1.00) |
| 1125016 | F374, S375, T376, F377, C379, F392, D427, E516 | 374-516 | 100% | 1 | 1 | 1 | 0 | 1.00 (0.04:1.00) |
| 1087820 | D428, F429, E516 | 428-516 | 100% | 1 | 1 | 1 | 0 | 1.00 (0.04:1.00) |
| 1087822 | T500, N501, G502 | 500-502 | 100% | 1 | 1 | 1 | 0 | 1.00 (0.04:1.00) |

Results Returned: 6 | Displaying: 6 Export Results

#### EPITOPE SUMMARY

T500, N501, G502 is a discontinuous epitope (epitope ID 1087822) studied as part of Spike glycoprotein from SARS-CoV2., tested in 7 B cell assays

#### COMPILED DATA

**B Cell Assay(s) 7**

| Assay Type | Positive / All |
|---|---|
| qualitative binding | 2 / 2 |
| neutralization | 1 / 2 |
| dissociation constant KD | 1 / 1 |
| off rate | 1 / 1 |
| on rate | 1 / 1 |

http://www.iedb.org/result_v3.php?cookie_id=6e2edb

6XC2 : Crystal structure of SARS-CoV-2 receptor binding domain in complex with neutralizing antibody CC12.1
Yuan, M., Liu, H., Wu, N.C., Zhu, X., Wilson, I.A. (2020) Science **369**: 1119-1123
**Released** 2020-07-08 **Method** X-RAY DIFFRACTION 3.112 Å
**Organisms** Homo sapiens SARS-CoV-2
**Macromolecule** CC12.1 heavy chain (protein) CC12.1 light chain (protein) Spike protein S1 (protein)

6XC4 : Crystal structure of SARS-CoV-2 receptor binding domain in complex with neutralizing antibody CC12.3
Yuan, M., Liu, H., Wu, N.C., Zhu, X., Wilson, I.A. (2020) Science **369**: 1119-1123
**Released** 2020-07-08 **Method** X-RAY DIFFRACTION 3.112 Å
**Organisms** Homo sapiens SARS-CoV-2
**Macromolecule** CC12.3 heavy chain (protein) CC12.3 light chain (protein) Spike protein S1 (protein)

7JMP : Crystal structure of SARS-CoV-2 receptor binding domain in complex with neutralizing antibody COVA2-39
Wu, N.C., Yuan, M., Liu, H., Zhu, X., Wilson, I.A. (2020) Biorxiv
**Released** 2020-08-26 **Method** X-RAY DIFFRACTION 1.712 Å
**Organisms** Homo sapiens SARS-CoV-2
**Macromolecule** COVA2-39 heavy chain (protein) COVA2-39 light chain (protein) Spike protein S1 (protein)

7JMO : Crystal structure of SARS-CoV-2 receptor binding domain in complex with neutralizing antibody COVA2-04
Wu, N.C., Yuan, M., Liu, H., Zhu, X., Wilson, I.A. (2020) Biorxiv
**Released** 2020-08-26 **Method** X-RAY DIFFRACTION 1.712 Å
**Organisms** Homo sapiens SARS-CoV-2
**Macromolecule** COVA2-04 heavy chain (protein) COVA2-04 light chain (protein) Spike protein S1 (protein)

6XKQ : Crystal structure of SARS-CoV-2 receptor binding domain in complex with neutralizing antibody CV07-250
Yuan, M., Liu, H., Zhu, X., Wu, N.C., Wilson, I.A. (2020) Cell
**Released** 2020-08-26 **Method** X-RAY DIFFRACTION 2.55 Å
**Organisms** Homo sapiens SARS-CoV-2
**Macromolecule** CV07-250 heavy chain (protein) CV07-250 light chain (protein) Spike protein S1 (protein)

6XKP : Crystal structure of SARS-CoV-2 receptor binding domain in complex with neutralizing antibody CV07-270
Liu, H., Yuan, M., Zhu, X., Wu, N.C., Wilson, I.A. (2020) Cell
**Released** 2020-08-26 **Method** X-RAY DIFFRACTION 2.72 Å
**Organisms** Homo sapiens SARS-CoV-2
**Macromolecule** CV07-270 heavy chain (protein) CV07-270 light chain (protein) Spike protein S1 (protein)

**6XGD:** https://www.rcsb.org/structure/6XDG Complex of SARS-CoV-2 receptor binding domain with the Fab fragments of two neutralizing antibodies **REGN-COV2**
**Released:** 2020-06-24 **Method:** ELECTRON MICROSCOPY **Resolution:** 3.90 Å
**Organism(s):** Severe acute respiratory syndrome coronavirus 2, Homo sapiens
**DOI:** 10.2210/pdb6XDG/pdb **EMDataResource:** EMD-22137
